# Rostral Associations of MRI Atrophy of the Amygdala and Entorhinal Cortex Across the AD Spectrum

**DOI:** 10.64898/2026.01.27.26344987

**Authors:** Michael I. Miller, Yi Xie, Kaitlin M. Stouffer, Can Ceritoglu, Jiabei Li, J. Tilak Ratnanather, Laurent Younes, Arnold Bakker, Nisha Rani, Marilyn S. Albert, Juan C Troncoso, Meaghan Morris, the Alzheimer’s Disease Neuroimaging Initiative

## Abstract

This paper examines associations of atrophy in the amygdala, entorhinal cortex and hippocampus based on magnetic resonance imaging (MRI) and Positron Emission Tomography (PET) scans from two independent cohorts: Alzheimer’s Disease Neuroimaging Initiative (ADNI) and Biomarkers of Cognitive Decline Among Normal Individuals (BIOCARD) study. The amygdala and entorhinal cortex (ERC) are shown to change earlier in the disease than the hippocampus based on atrophy of laminar thickness of the ERC and amygdala volumes. Over four hundred laminar reconstructions showed that ERC volume loss is linked to cortical thinning, as a more specific measure historically linked to the layer specific pattern of tau pathology deposition. Additionally, high field atlasing with delineations of amygdala subregions shows predominant volume loss in medial subregions including basomedial, basolateral, and corticocentromedial compared with the lateral subregion. In the context of earlier work linking MRI-based atrophy with hyperphosphorylated tau deposition in the ERC and amygdala, the atrophy rate marker is shown to be strongly associated with tau deposition as measured by tau positron emission tomography imaging and co-localization of the atrophy marker to the spatial distribution of tau deposition.

**Highlights:**

- Structural MRI analyses across ADNI and BIOCARD cohorts reveal significantly greater early atrophy in amygdala and entorhinal cortex (ERC), marking them as sensitive indicators of preclinical Alzheimer’s disease.
- ERC volume loss is shown to correspond to cortical laminar thinning by surface-based diffeomorphic reconstructions, confirming MRI volume atrophy as a biologically valid marker of early neuronal degeneration.
- Predominant atrophy in medial amygdala subregions (basomedial, basolateral, corticocentromedial) are identified compared to the lateral amygdala using high-field (11T) atlases, mirroring known histopathological tau distribution.
- MRI volume atrophy correlates with tau burden measured by tau PET imaging, demonstrating region-specific correspondence between structural atrophy and molecular pathology.

## 1. Introduction

Alzheimer’s disease (AD) remains the leading cause of dementia all over the world. With new clinical therapies and trials targeting the pathology underlying AD, early diagnosis through biomarkers becomes increasingly important. AD neuropathological changes begin many years before clinical symptoms, leading to a new framework for classifying AD pre-symptomatic stages using biomarkers of AD pathology [1, 2]. Recent progress in neuroimaging offers opportunity to localize structural and molecular changes in vivo reflecting underlying AD pathologies to improve early detection and diagnostic accuracy for mild cognitive impairment (MCI) and AD dementia.

The medial temporal lobe (MTL) remains especially important in the classic neuropathological staging of early AD. Supratentorial tau pathology first arises in the transentorhinal cortex (TEC) (sometimes considered part of the perirhinal cortex) and adjacent entorhinal cortex (ERC), subsequently extending to involve the hippocampus, before ultimately spreading throughout the neocortex as described by Braak and Braak [3], This pathological sequence is the basis for the Braak staging of AD tau pathology from I to VI [3, 4] and cornerstone of the current system to determine the severity of Alzheimer’s disease neuropathological change (ADNC) [5]. The description of this pathological sequence weighs heavily on the early involvement of the ERC followed by hippocampus in stages I or II, but does not focus on the amygdala until stage III [3, 4]. Following Braak’s landmark observations, histological studies in early clinical stages of AD have targeted predominantly the ERC/TEC, e.g., the loss of neuronal patterns in layer II therein, even in the mildest stages of AD [6]. However, other investigators venturing beyond the ERC/TEC found that other limbic regions such as the amygdala also accumulate pathology relatively early in the disease [7, 8, 9, 10, 11], with tau pathology in the amygdala in AD distributed in specific spatial patterns [8, 9, 12].

These observations, based on postmortem examinations, are now being complemented by emerging neuroimaging findings of very early atrophy of the amygdala on structural magnetic resonance imaging (MRI) and tau lesions by Positron Emission Tomography (PET) many years prior to the onset of MCI symptom [13, 14, 15, 16]. Likewise, recent tau PET imaging studies show tau pathology in amygdala early in the disease course [17, 18, 19, 20, 21, 22]. This early involvement of the amygdala is consistent with its strong interconnections with the entorhinal-hippocampal network and has gained increasing attention in recent research.

Neuronal loss and synaptic injury in AD presumably lead to the measurable atrophy (volume loss) on structural MRI scans that we and others are demonstrating in the ERC and amygdala. In the Alzheimer’s Disease Neuroimaging Initiative (ADNI) cohort, Kulason et al. demonstrated that atrophy of the entorhinal cortex in MRI correlated with clinical severity in AD – patients with smaller ERC volumes in fact tended to have worse memory performance and more advanced symptoms [23]. Longitudinal MRI studies further demonstrate that ERC atrophy can precede and predict cognitive decline [24]. Importantly, a changepoint analysis demonstrated that the accelerations in atrophy (i.e., changepoints) occur almost 10 years prior to MCI symptom onset, many years preceding the changepoint for the hippocampus [25].

MRI studies targeting the amygdala have been more limited, but evidence indicates the amygdala undergoes early atrophy [26]. These studies show decreased amygdala volume correlating with decreased memory/cognition [27, 28, 29, 30, 31, 32] and increased severity of neuropsychiatric symptoms [33, 32]. A recent study by our group using ADNI data showed significantly higher rates of amygdalar atrophy rates for cognitively unimpaired individuals who progressed to MCI than cognitively unimpaired individuals who remained stable [12].

Recent developments of high-resolution MRI atlases and deep learning segmentation now allow more accurate labeling of MTL substructures. Previously, reliable MTL segmentation has been challenging due to boundary indistinction and tracing variation. In the case of ERC, its proximity to the cerebrospinal fluid in the collateral sulcus and to the optic chiasm can confound automated segmentation algorithms given its thin laminar structure.

Moreover, the use of a Large Deformation Diffeomorphic Metric Mapping (LDDMM) framework enables the computation of thickness atrophy rate in the laminar structures of the ERC far more accurately than before [34], allowing us to demonstrate that the volume changes we have seen early in the entorhinal regions are likely associated with laminar cortex thinning. This is important given the apparent loss of neuronal patterns in layer II associated with early disease. Simultaneously, with the advent of our high field atlases, we have been able to define high-field 11T templates which partition the amygdala into subregions, allowing us to examine the locality the morphometry marker to regions including basomedial amygdala (BMA), basolateral amygdala (BLA), corticocentromedial amygdala (CMA), and lateral amygdala (LA). By employing the high-resolution diffeomorphic mapping of amygdalar subregions, we can explore the sub-nuclear atrophy patterns within cohorts – seeking to determine if the selective vulnerability of the medial amygdala observed is recapitulated in our sample.

This paper examines data from two independent cohorts, the Alzheimer’s Disease Neuroimaging Initiative (ADNI) study and the Biomarkers of Cognitive Decline Among Normal Individuals (BIOCARD) study, which include data from cognitively unimpaired individuals, those with MCI and those with mild AD dementia. The questions that we examine are four-fold: (1) Do the ERC and amygdala in both samples demonstrate greater atrophy than the hippocampus early in the course of AD, (2) Is the atrophy of ERC volume linked to cortical thinning, as may be implied by the loss of layer II neurons in early disease, (3) Given progress in high field imaging and sub-parcellations, can the earliest morphometric changes occurring in the amygdala be tied to its local neighborhood geometry and presumed circuitry with the entorhinal cortex, and (4) Given the stability of the MRI-based atrophy marker, and prior evidence by our group that tau deposition in brain tissue is associated with MTL atrophy [12], can we demonstrate an association between the MRI atrophy markers and tau burden, as measured by PET. These efforts are intended to advance the development of sensitive and specific imaging biomarkers for the preclinical and prodromal stages of AD, when therapeutic interventions may be most effective.

## 2. Material and Methods

Data were obtained from the BIOCARD study and the ADNI database. The BIOCARD Study (www.biocard-se.org) was established in 1995. It was originally led by investigators at the National Institutes of Health (NIH) and was transferred to the research team at the Johns Hopkins School of Medicine in 2009. It is supported by funding from the National Institute on Aging and is led by Dr. Marilyn Albert. The overarching goal of the study is to improve understanding of the earliest phases of AD, accelerate the development of new treatments that are directed at the basic mechanisms of the disease and hasten the time when effective treatments for AD and related disorders become a reality. ADNI (https://adni.loni.usc.edu) was launched in 2003 as a public-private partnership, led by Principal Investigator Michael W. Weiner, MD. The primary goal of ADNI has been to test whether serial MRI, PET, other biological markers, and clinical and neuropsychological assessment can be combined to measure the progression of mild cognitive impairment (MCI) and early AD.

### 2.1. BIOCARD Participants and Imaging Data

Participants in BIOCARD study were categorized into four diagnostic groups based on longitudinal clinical evaluations: cognitively normal (CN), impaired not MCI, MCI, and AD. Subjects in all groups were required to have at least three 3T MR scans for longitudinal comparison. Demographic summaries, including baseline age, scan count, and follow-up duration, are provided in Table 2. All MRI data were acquired on a 3T MRI system using a 32-channel head coil (Achieva, Philips Healthcare, Best, The Netherlands). A 3D T1-weighted Magnetization-Prepared-Rapid Gradient-Echo (MPRAGE) image was acquired with the following parameters: TR = 6.5 ms, TI = 843 ms, TE = 3.11 ms, turbo factor = 240, shot interval = 3000 ms, flip angle = 8^◦^, voxel size = 1 × 1 × 1 mm^3^, FOV = 240 × 256 × 204 mm^3^, and scan duration = 5 min 59 s. Longitudinal 1.5 T structural MRI data acquired with both sagittal and coronal slice orientations were additionally used to validate the robustness of the estimated atrophy-rate patterns. Sagittal acquisitions had a voxel resolution of 1.5 0. × 9375 × 0.9375 mm^3^, while coronal acquisitions had a voxel resolution of 0.9375 × 2.0 0 ×.9375 mm^3^. Detailed acquisition parameters for these scans have been previously reported [25].

**Table 1:** Statistics of training data and manual segmentations for ADNI and BIOCARD; nnU-Net was trained for the different datasets.

|  | **ADNI** | **BIOCARD** |
| --- | --- | --- |
| # Scans | 294 | 371 |
| # Subjects | 67 | 107 |
| # of time points per subject | 4.39 | 3.47 |

**Table 2:**
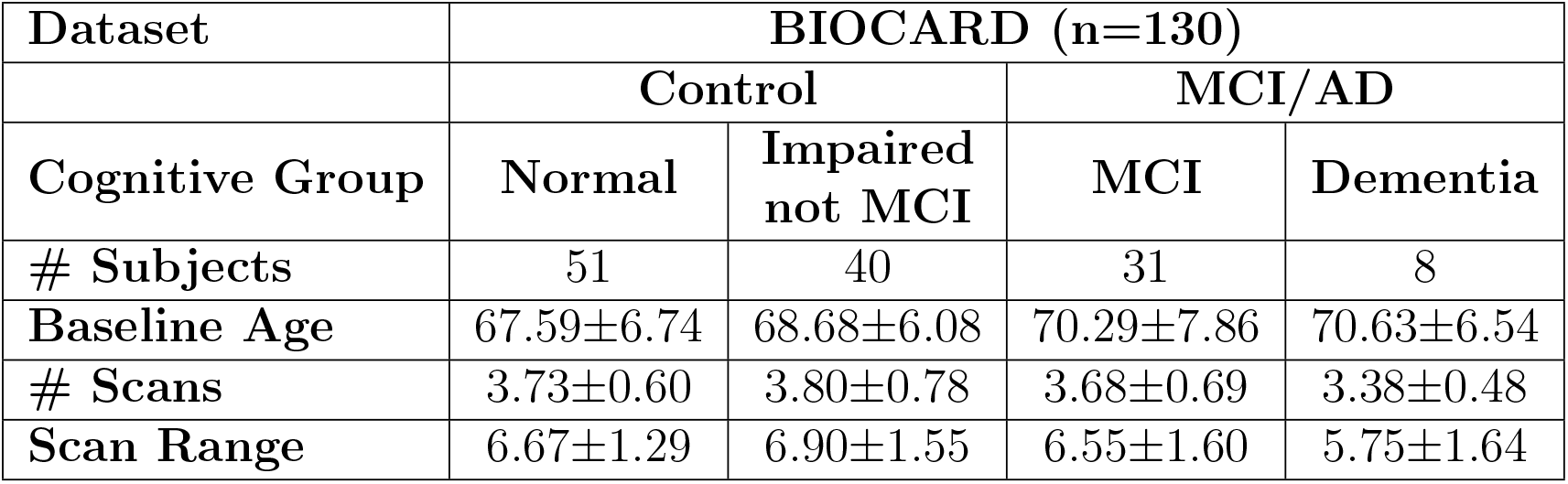
Demographics of four subgroups of BIOCARD dataset. Statistics reported as mean ± standard deviation.

PET scans were acquired on a GE DISCOVERY RX PET/CT scanner in three-dimensional acquisition mode. For tau imaging, ^18^F-MK6240 PET was performed 90 ± 1min after intravenous injection of 5.0 ± 10% mCi of tracer, followed by a saline flush, yielding six 5-min frames. For amyloid-*β* imaging, ^11^C-Pittsburgh compound-B (^11^C-PiB) PET was obtained 40 min post-injection of 15 ± 1.5 mCi, reconstructed into four 5-min frames. Residual syringe activity was measured to ensure accurate dose quantification. Image reconstruction used the 3D ordinary Poisson ordered-subset expectation-maximization (3D OP-OSEM) algorithm with corrections for detector efficiency, decay, dead time, attenuation, and scatter (Hudson and Larkin, 1994). Attenuation correction employed low-dose CT (120 keV, 80 mA, slice thickness = 3.75 mm, separation = 3.3 mm, FOV = 500 mm) reconstructed with the standard kernel. Final PET volumes consisted of 2 × 2 × 3.27 mm^3^ voxels representing radioactivity concentration (Bq/mL).

### 2.2. ADNI Participants and Imaging Data

Diagnostic categories of ADNI in stage 1,2 and GO included CN, SMC (Subjective Memory Concern), MCI, and LMCI (Late Mild Cognitive Impairment), as shown in Table 3. Subjects in these stages were required to have an anteriorly continuous collateral sulcus (CS), as detailed [23]. Diagnostic categories of ADNI in stage 3 and 4 included CN, SMC, EMCI (Early Mild Cognitive Impairment), MCI, LMCI and AD, and subjects in these stages were required to have at least three 3T MR scans for longitudinal comparison, with demographic data shown in Table 4. T1-weighted MRI images were acquired across multiple sites on GE, Philips, and Siemens scanners at 3.0 T. Sequences typically included 3D MPRAGE and 3D IR-FSPGR (BRAVO). Acquisition parameters varied across protocols, with repetition times (TR) typically 2300 ms (Gradient TR 6.8 ms for Philips), echo times (TE) from 2.8 to 4.1 ms, inversion times (TI) 400/900/1000 ms when applicable, and flip angles 8^◦^–11^◦^. Imaging was performed with voxel dimensions of 1.2 × 1 × 1 mm^3^, fields of view (FOV) 240*/*256 × 256 × 176–211 mm^3^. All acquisitions employed multi-channel head coils (8–32 channels depending on scanner model).

**Table 3:**
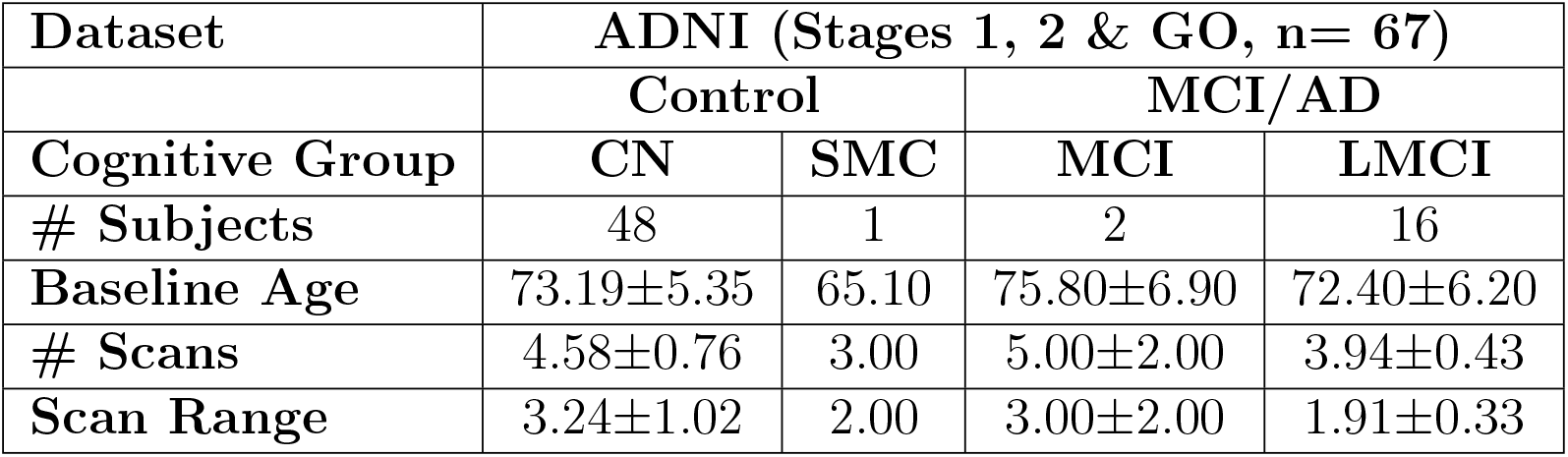
Demographics of ADNI 1, 2 & GO datasets. Statistics reported as mean ± standard deviation.

**Table 4:** Demographics of ADNI 3 & 4 datasets. Statistics reported as mean ± standard deviation.

| Dataset | ADNI (Stage 3 & 4) |  |  |  |
| --- | --- | --- | --- | --- |
| Control | CN |  | SMC |  |
| # Subjects | 174 |  | 8 |  |
| Baseline Age | 72.94±7.02 | | 74.25±6.34 | |
| # Scans | 3.54±0.75 | | 3.00±0.00 | |
| Scan Range | 5.17±1.67 | | 2.05±0.17 | |
| MCI/AD | EMCI | MCI | LMCI | AD |
| # Subjects | 24 | 103 | 10 | 19 |
| Baseline Age | 75.75±7.01 | 73.24±7.53 | 74.30±7.47 | 74.26±8.52 |
| # Scans | 3.88±0.93 | 3.97±0.98 | 3.60±0.66 | 3.20±0.54 |
| Scan Range | 3.64±1.11 | 4.48±1.75 | 3.17±1.10 | 2.81±1.19 |

PET imaging was conducted using ADNI-qualified PET/CT scanners including GE, Philips, and Siemens systems in three-dimensional acquisition mode. Amyloid-*β* PET imaging was conducted with [^18^F]AV-45 (Florbetapir), administrated as 370 ± 10% (10.0 ± 1.0 mCi) MBq intravenous bolus, with imaging beginning 50 min post-injection and reconstructed into four 5-minute frame. Tau PET imaging was conducted with [^18^F]AV-1451 (Flortaucipir), administered as a 370 ± 10% (10. ± 0 1.0 mCi) MBq intravenous bolus, with imaging beginning 75 min post-injection, reconstructed into six 5-minute frames. All radiotracer injections were followed by a 0.9% sterile sodium chloride flush, and residual syringe activity was recorded for accurate quantification of injected dose. Attenuation correction was performed using low-dose CT scans, typically four 5-min frames for Florbetapir and Florbetaben, and six 5-min frames for AV-1451. Reconstruction parameters vary among different scanners, typically using OSEM-based algorithms [35]. Final PET voxel values represent the scanner-reconstructed activity concentration, typically proportional to radioactivity concentration (Bq/mL).

### 2.3. Data preprocessing

All T1-weighted structural MRI scans and PET datasets were processed using a standardized pipeline. For each subject, raw DICOM directories were scanned to identify all available PET acquisitions and classify them as either amyloid or tau. T1-weighted MPRAGE MRI scans were likewise identified and converted from DICOM to NIfTI format using the dcm2niix conversion software [36]. PET series were converted to NIfTI format according to their acquisition protocol, resulting in either 4 × 5-minute or 6 × 5-minute dynamic 4D volumes.

Dynamic PET data were preprocessed using SPM12 [37]. All temporal frames were realigned to the first frame to correct for intra-scan motion, and a static PET volume was generated by simple unweighted averaging of the motion-corrected frames. Each mean PET volume was paired with the temporally closest T1-weighted MPRAGE MRI based on acquisition dates. Rigid-body PET-to-MRI coregistration was performed using FSL FLIRT [38]. Registration quality was verified through visual inspection of PET–MRI overlays in all three orthogonal planes, and datasets showing inadequate alignment were excluded or reprocessed using FSL tools in place of SPM when necessary.

T1-weighted MPRAGE images were additionally processed for skull stripping using DeepBrain, an open-source TensorFlow-based brain-extraction toolkit [39]. DeepBrain applies a convolutional neural network to generate a brain-only mask that removes scalp, skull, and other non-brain tissues. All brain-extracted images were visually inspected to ensure accurate preservation of cortical and subcortical anatomy prior to subsequent segmentation.

### 2.4. MTL segmentation and atrophy rate calculations

Automated segmentation of five medial temporal lobe (MTL) structures – amygdala, ERC, TEC, anterior hippocampus (HA), and posterior hippocampus (HP) – was performed on T1-weighted 3T MRI using a combination of deep learning and atlas-based methods. Specifically, ASHS [40] was used to delineate HA and HP subfields. Combined with manual segmentations of amygdala, ERC, and TEC, such delineations served as ground truth for training.

Segmented images were used in developing a self-configuring deep convolutional neural network called nnU-Net [41]. nnU-Net automatically configures the entire segmentation pipeline — including preprocessing, network architecture, training schedule, data augmentation, and postprocessing — based solely on the properties of the training dataset. It builds upon a 3D U-Net backbone but adapts key hyperparameters and architectural components to match the data characteristics, enabling state-of-the-art performance without manual tuning. The model was separately trained on 294 ADNI scans and 371 BIOCARD scans (Table 1), with an 80:20 train/test split and 5-fold cross-validation to assess performance. All models incorporated focal loss to address class imbalance. A collage of the training data with corresponding manual segmentations superimposed is shown in Figure 1.

**Figure 1.**
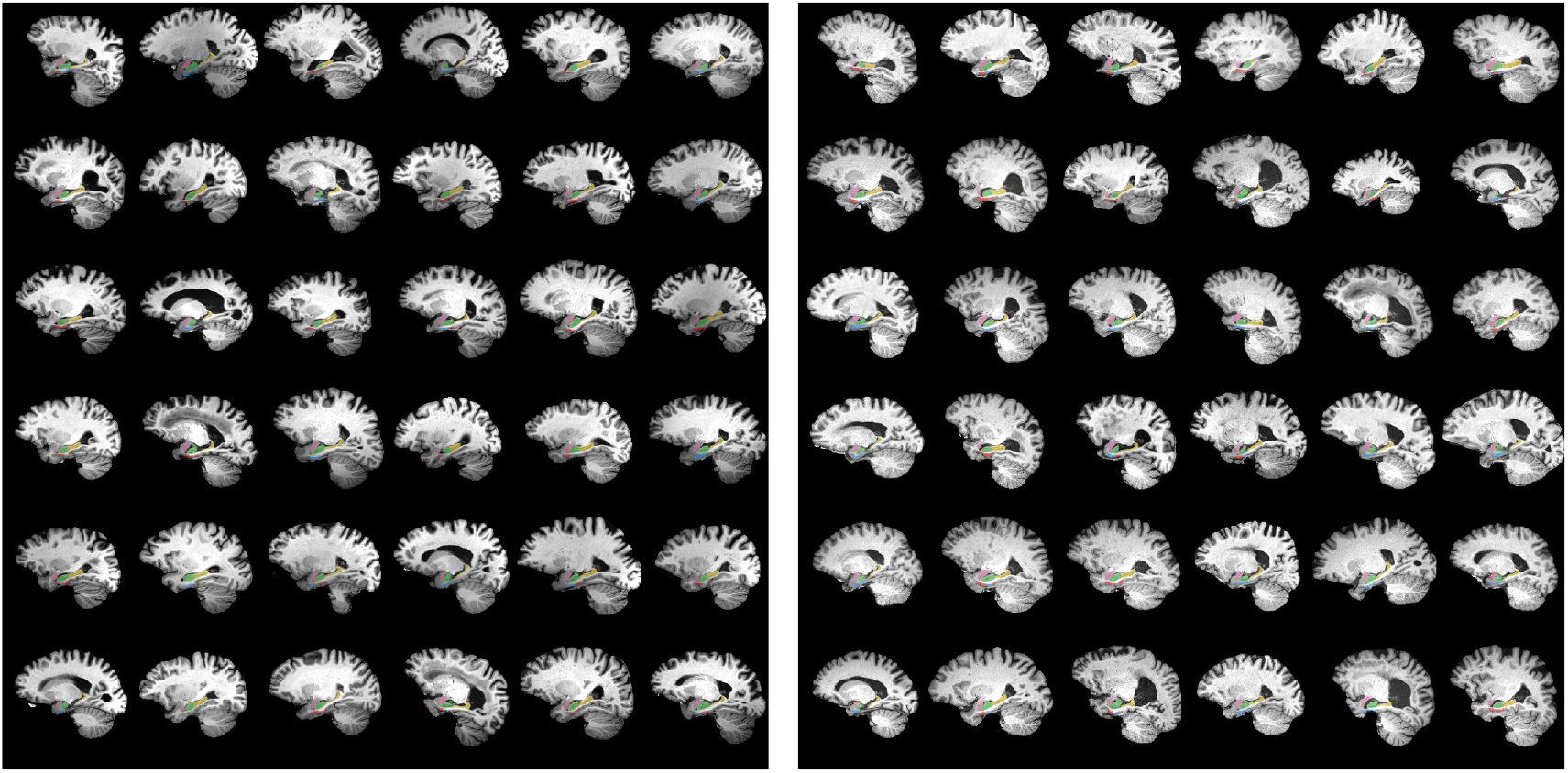
Sagittal sections from fully 3D training data for the nnU-Net manual segmentatiom of 294 and 371 ADNI (left) and BIOCARD (right) T1 scans. Shown are subsets of the segmentations which are overlaid with opacity of 40%, including amygdala (pink), ERC (blue), TEC (green), hippocampus anterior (green) and hippocampus posterior (yellow).

Atrophy rates were calculated for each subject using longitudinal volume estimates derived from region-specific segmentation masks. Volumes were computed by multiplying the number of voxels within each label by the voxel dimensions (1.0 × 1.0 × 1.2 mm). A linear regression model was fitted to each subject’s longitudinal volume trajectory to estimate the annualized rate of change, expressed as both absolute volume change (mm^3^/year) and percent change relative to baseline. Standard deviation of all predicted segmentation in each region (amygdala, ERC/TEC, hippocampus) is calculated, and time points whose regions exceed 2.5 the standard deviation from the group mean within the subject were labeled as outliers and excluded from subsequent analysis. Visual inspection was also performed to remove outliers due to poor image quality.

### 2.5. LDDMM Framework for diffeomorphic registration

LDDMM models transformation as geodesic flows on the manifold of diffeomorphisms, and guarantees smooth, invertible mappings that preserve topological structures of anatomy [42]. Let *I*_0_ and *I*_1_ : ℝ^3^ → ℝ denote the source and target images, respectively. LDDMM seeks a time-dependent velocity field *v*(*x, t*) on a suitable smoothness space *V* (a reproducing kernel Hilbert space, RKHS) that minimizes a variational energy:

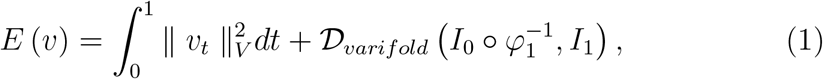

Where 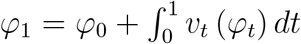, with *φ*_0_ (*x*) = *x* is the resulting diffeomorphic warp at 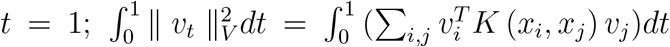 is a smoothness regularizer using RKHS; 𝒟_*varifold*_(·, ·) is a data fidelity between deformed source 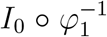 and *I*_1_ via a varifold-based shape dissimilarity metric that compares the distribution of labeled surface/region elements. The optimal solution of this variational problem corresponds to a geodesic path in the diffeomorphism group connecting *I*_0_ to *I*_1_. Our pipeline for LDDMM of the population is depicted in Figure 2. We project onto all of the segmented structures of the MTL the high field atlas published in [12].

**Figure 2.**
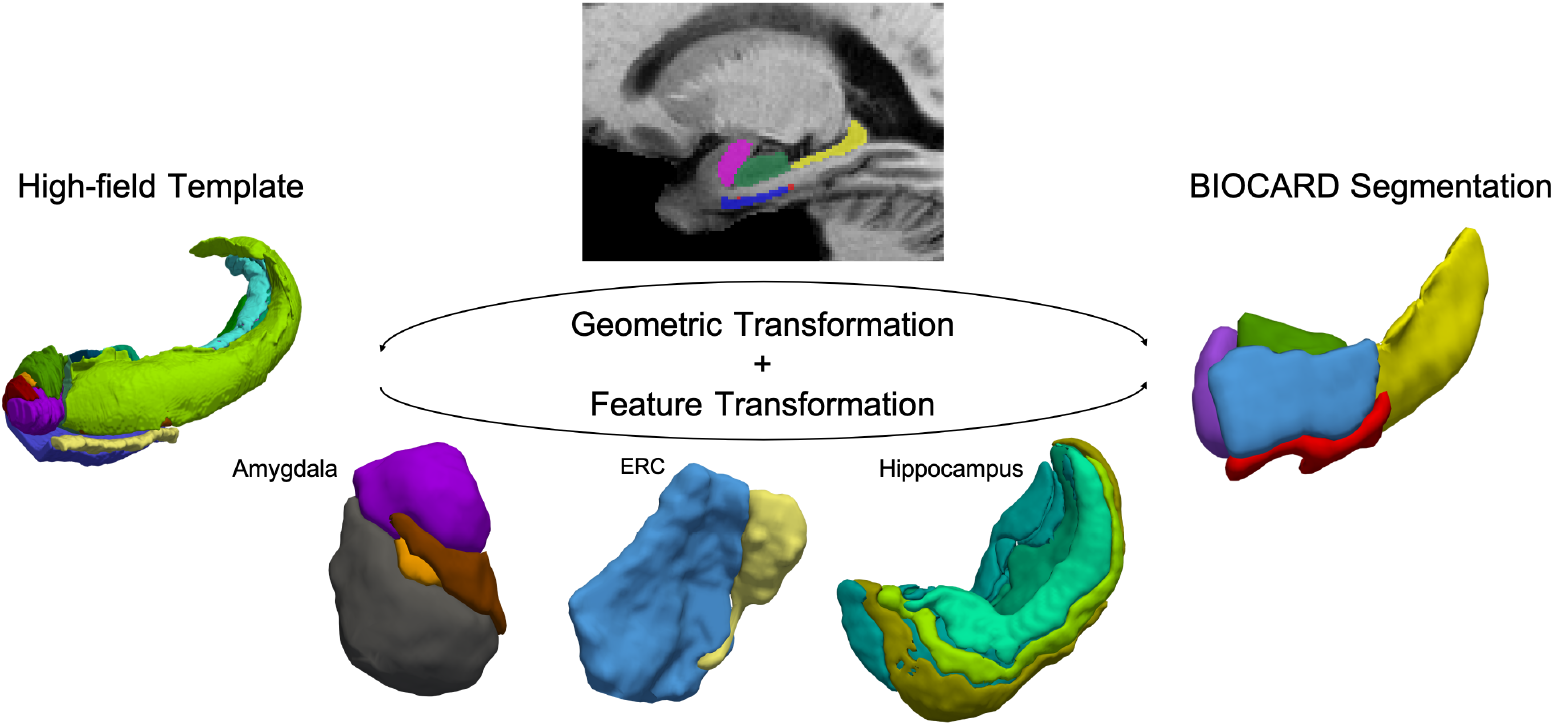
Diffeomorphic mapping from high-field template to BIOCARD and ADNI segmentations depicting both geometric transformation and transfer of template labels.

In addition, to examine the atrophy rates within the amygdala, we used high-field 11T MRI atlases parcellated into up to 5 subregions, where present, based on known anatomical markers [43, 44, 45] that could be linked to visible differences in MR intensity [12]. The subregions included basomedial amygdala (BMA), basolateral amygdala (BLA), corticocentromedial amygdala (CMA), lateral amygdala (LA), and periamygdaloid area (PA). Briefly, subregions were delineated on coronal slices posterior to anterior, with the posterior boundary of the amygdala comprised of CMA, and moving anterolaterally, the posterior boundaries of BMA, BLA, and LA respectively defined. PA was differentiated specifically from the cortical region of CMA by the presence of laminated grey levels. LA typically extended to the anterior most part of the amygdala, with dark contrast lines likely representing amygdalafugal pathways seen in the anterior dorsal region.

To compute volumetric atrophy within amygdala subregions, we employed a two-stage diffeomorphic mapping procedure from the high-field template to each subject’s 3T segmentation. For each subject, all available time points were jointly aligned in a global LDDMM space using particle based varifold LDDMM [46] from the amygdala, ERC, TEC, and hippocampus. A subject-specific midpoint template was then generated by computing the average initial momentum across time points and deforming the high field atlas. This subject-specific template was subsequently mapped to each individual time point using volume LDDMM [42].

Following deformation, voxel-wise segmentation masks were reconstructed from the particle representation by interpolating particle-defined anatomical boundaries onto a regular grid and assigning each voxel to the subregion with the highest empirical likelihood. Atrophy rates were computed from the slope of the fitted longitudinal model to the structures.

### 2.6. Entorhinal Cortex Reconstruction and Cortical Thickness Calculation

To quantify cortical thickness within the ERC and TEC, we employed a surface-based diffeomorphic framework inspired by prior work in longitudinal shape analysis of the medial temporal lobe. Figure 3 depicts our pipeline for cortical thickness mapping [34]. We estimated vertex-wise cortical thickness by computing the geodesic flow between paired cortical surfaces: the inner (white matter) and outer (pial) boundaries applying LDDMM to register the inner surface to the outer surface with the additional constraint that the deformation is restricted to the surface normal direction. We used the lengths of the flow lines, provided by each vertex trajectory under the transformation, to define the vertex-wise thickness between the cortical boundaries. The median of the distribution of the vertex-wise thickness was used to attribute a scalar thickness measurement to each ERC/TEC volume. The bottom left panel shows the thickness color scale across the ERC surface. The atrophy rate per subject was measured in the ERC/TEC as percentile change in thickness per time (% / year). Among all the generated surfaces, some have thin flagged TEC from the first time point, which vanished in later time points and caused inconsistency in thickness calculation. These subjects were not included in the calculation of volume-thickness atrophy correspondence.

**Figure 3.**
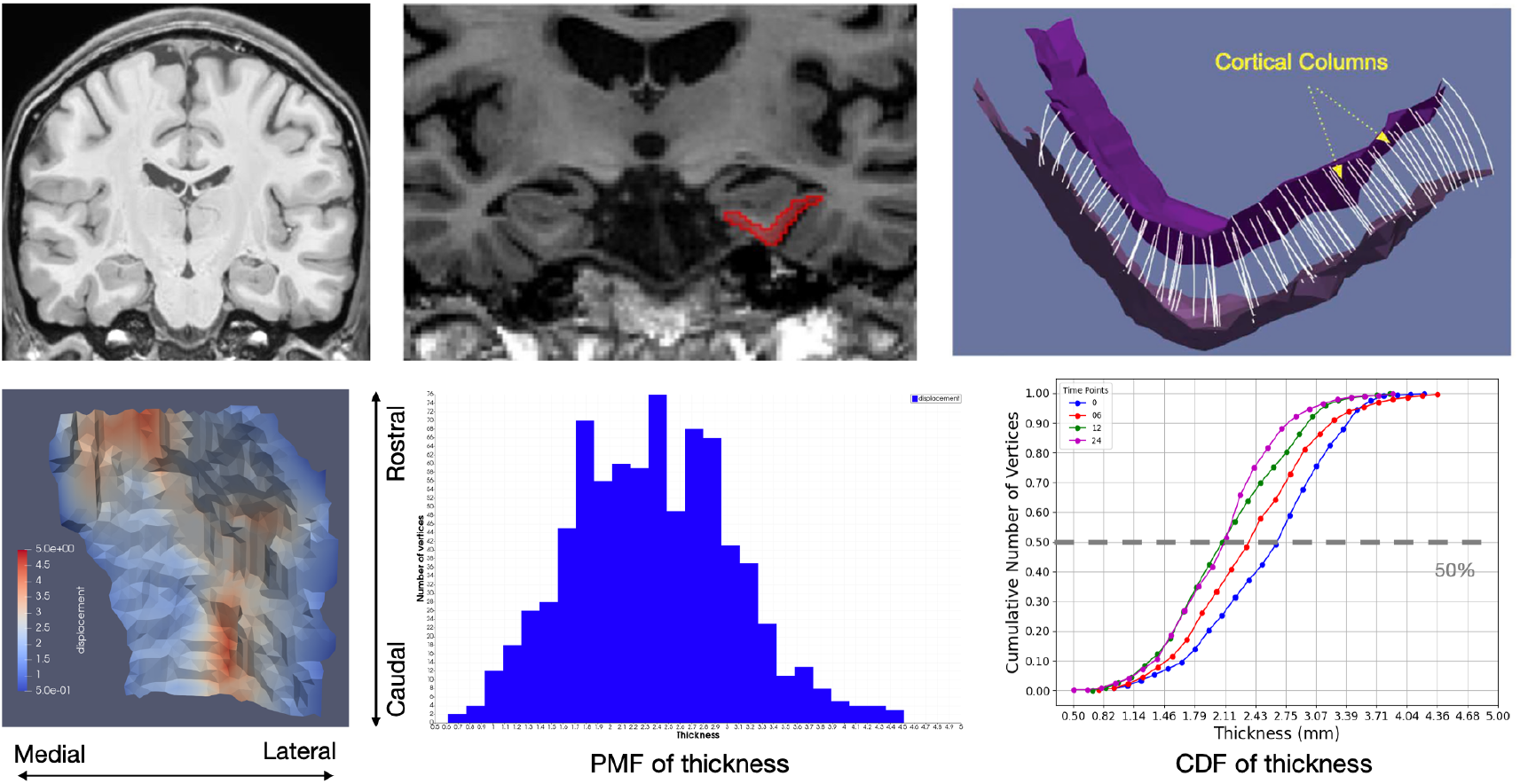
Top: pipeline for ERC reconstruction of thickness profiles. Bottom: thickness from 1-5 mm (left); thickness distribution (middle), CDFs of thickness loss (right).

We used the LDDMM triangulated mesh framework with hybrid elastic energy [47, 48, 49] to transport the thickness scalar field on the target surfaces onto the common coordinate system of a template estimated averaging all baseline outer surfaces [50].

### 2.7. PET accumulation of tau and amyloid tangles

For region-specific PET characteristics, we computed the Standardized Uptake Value Ratio (SUVR) for each region of interest (ROI) [51]. For each MTL ROI compartment the radioactivity concentration (Bg/mL) was calculated dividing the sum of voxel intensity within the ROI by its volume. This value was normalized by the reference tracer concentration averaged across the voxels of the cerebellar gray matter. The intensity ratio for an ROI is defined as:

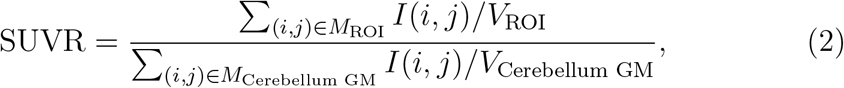

where *I*(*i, j*) denotes the PET intensity at voxel (*i, j*), *M* denote voxel ensembles of different regions, *V*_ROI_ and *V*_cerebellum GM_ represent the volumes of the ROI and cerebellum grey matter, respectively. Cerebellar gray matter segmentation was generated using MRICloud, an atlas-based automated brain segmentation platform [52]. The resulting SUVR provides a comparative measure of the signal intensity within a given anatomical structure relative to the non-MTL brain tissue.

## 3. Results

Data from a total of 130 BIOCARD study participants, 67 ADNI participants (stages 1, 2 & GO) and 338 ADNI participants (stages 3 & 4) were available for analysis. Tables 2, 3 and 4 provide details about the number of participants, based on diagnostic categories at follow-up, the number of MRI scans included in the analyses and the mean years of follow-up.

### 3.1. MRI segmentations

Representative examples of structural MRI imaging from participants in the BIOCARD and ADNI cohorts are shown in Figure 4. T1-weighted sagittal slices illustrate anatomical integrity and contrast in both datasets capturing regional accumulation of AD-related pathology. Structural consistency across both cohorts supported joint analysis of volumetric trajectories. The right column of Figure 4 shows 3D renderings of the segmentations predicted from the T1-weighted images using our pipeline. Surface meshes were reconstructed and smoothed using the ITK-SNAP toolkit [53]. Visualizations from both datasets demonstrate the segmentation algorithm’s ability to generate anatomically plausible labels in regions such as the amygdala, ERC, and hippocampus, serving as the basis for quantitative morphometry analyses.

**Figure 4.**
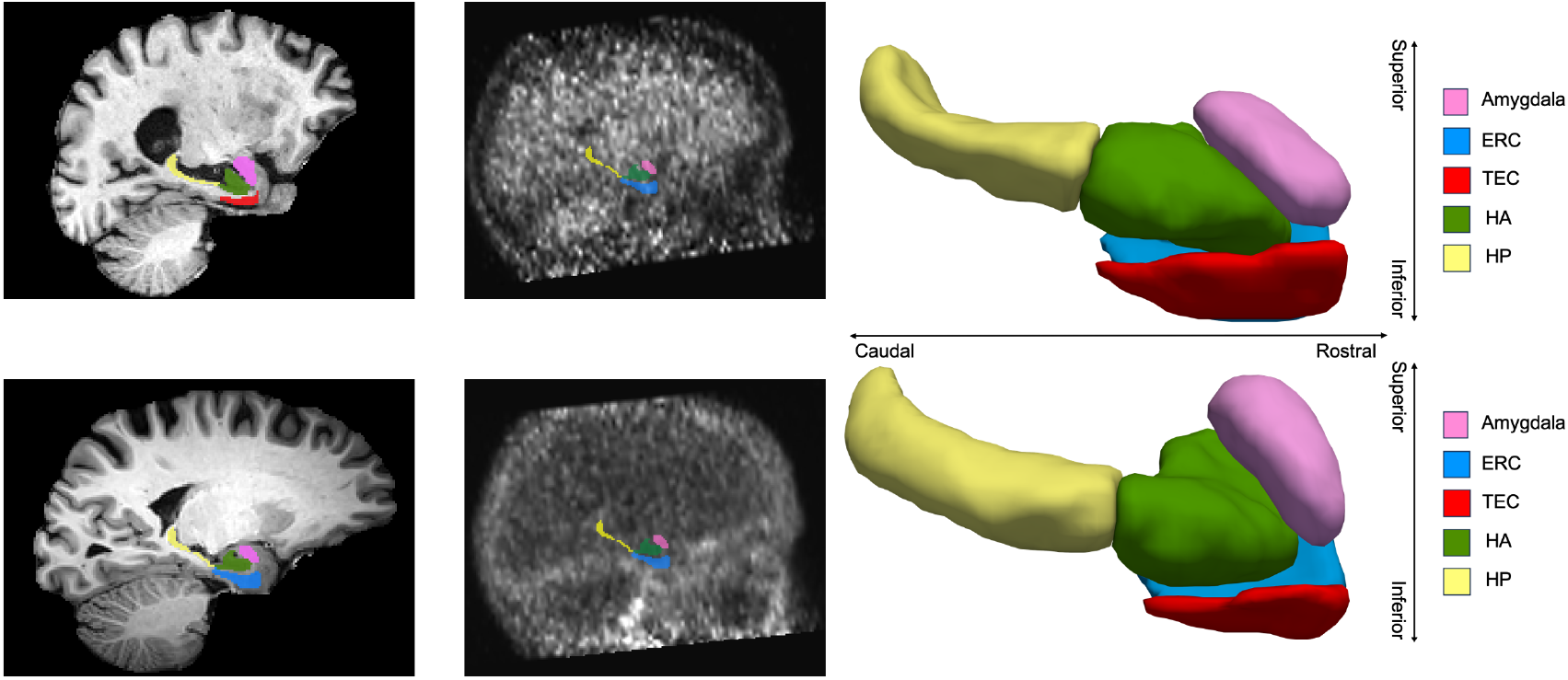
Left: sagittal slice of T1-weighted imaging for a subject in ADNI (top) and BIOCARD (bottom). Middle: tau PET and amyloid PET imaging for a subject in BIOCARD. Right: 3D segmentation of T1-weighted imaging for a subject in BIOCARD (top) and ADNI (bottom), smoothed by toolkit in ITK-SNAP.

### 3.2. Machine learning segmentations are consistent and accurate relative to manual raters and other methodologies

Figure 5 shows overlaps of manual and nnU-Net-predicted segmentation in BIOCARD (top two rows) and ADNI (bottom two rows). The dark colors denote overlapping voxels, and light colors denote differences between manual and predicted segmentations. Segmentation accuracy was quantitatively evaluated using Dice similarity coefficients, comparing nnU-Net predictions to manual segmentations. As visualized and summarized in Table 5, the classifier trained on hundreds of hand segmented MTLs exhibits high Dice scores across all five regions of interest (ROIs) in both the BIOCARD and ADNI datasets, with those within the ERC and TEC (0.87) similar to those reported for intra-rater reliability of manual segmentations in this area (0.85-0.88) [54]. Notably, segmentations of anterior and posterior hippocampus achieved Dice scores exceeding 0.95 in BIOCARD and above 0.92 in ADNI. The amygdala, ERC, and TEC exhibited slightly lower Dice scores, particularly in ADNI, where image contrast varied more substantially across subjects. This reduction likely reflects the geometric and cytoarchitectonic complexity of the ERC and TEC regions as laminar structures, which pose greater challenges for automated segmentation, as well as the reported variability across human architecture in these areas [54].

**Table 5:**
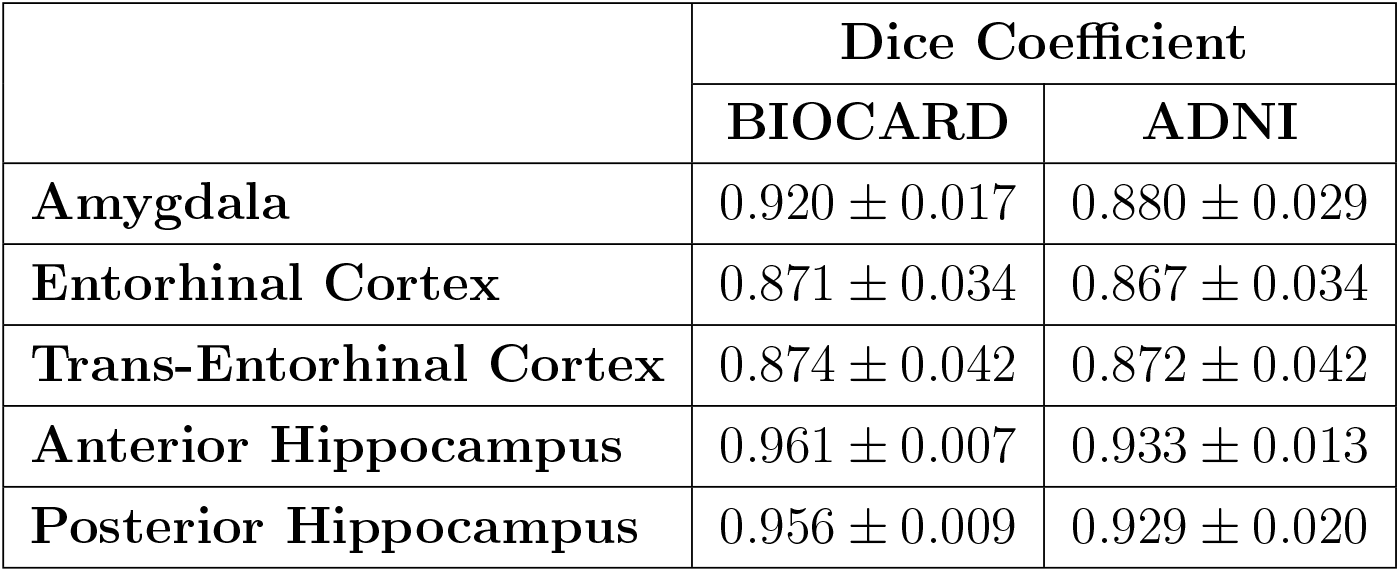
Dice coefficients of different structures in BIOCARD and ADNI based on nnU-Net classifier.

**Figure 5.**
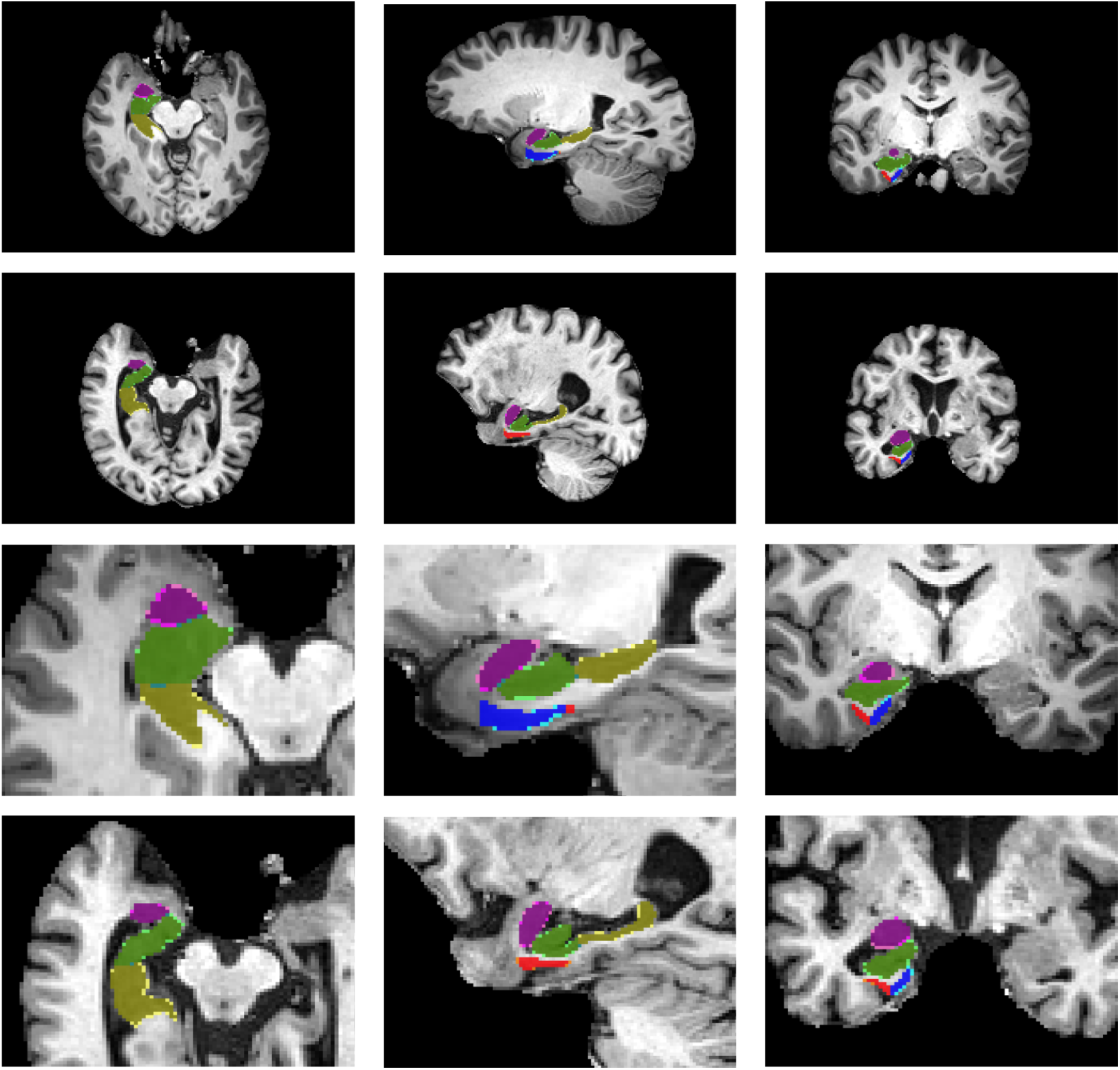
Overlap of manual and nnU-Net predicted segmentation in BIOCARD (1st and 3rd rows) and ADNI (2nd and 4th rows); dark colors denote overlaps and light colors denote differences.

We also compared our nnU-Net atrophy rates to other state-of-the-art methods including ASHS [40] for the hippocampus. We found that the atrophy rates obtained from the nnU-Net applied to the entire MTL gave almost identical atrophy rates. For the MCI group for BIOCARD, the atrophy rates were 0.6 and 0.65 % volume change per year with nnU-Net and ASHS, respectively; for ADNI 3.2% and 3.6%, respectively.

### 3.3. BIOCARD and ADNI predict ordered atrophy: ERC ∼ Amygdala > Hippocampus

Longitudinal MRI-based volume trajectories revealed a consistent ordering of regional atrophy rates across disease stages in both the BIOCARD and ADNI datasets. Figure 6 shows the trajectories of the MCI/AD subjects as evidenced by the atrophy rates of each individual ERC structure for all the subjects analyzed. The left and right columns correspond to the left and right hemispheres, respectively, and shows ERC/TEC atrophy rates for the BIOCARD MCI/AD sample (top row) and the ADNI sample (bottom row).

**Figure 6.**
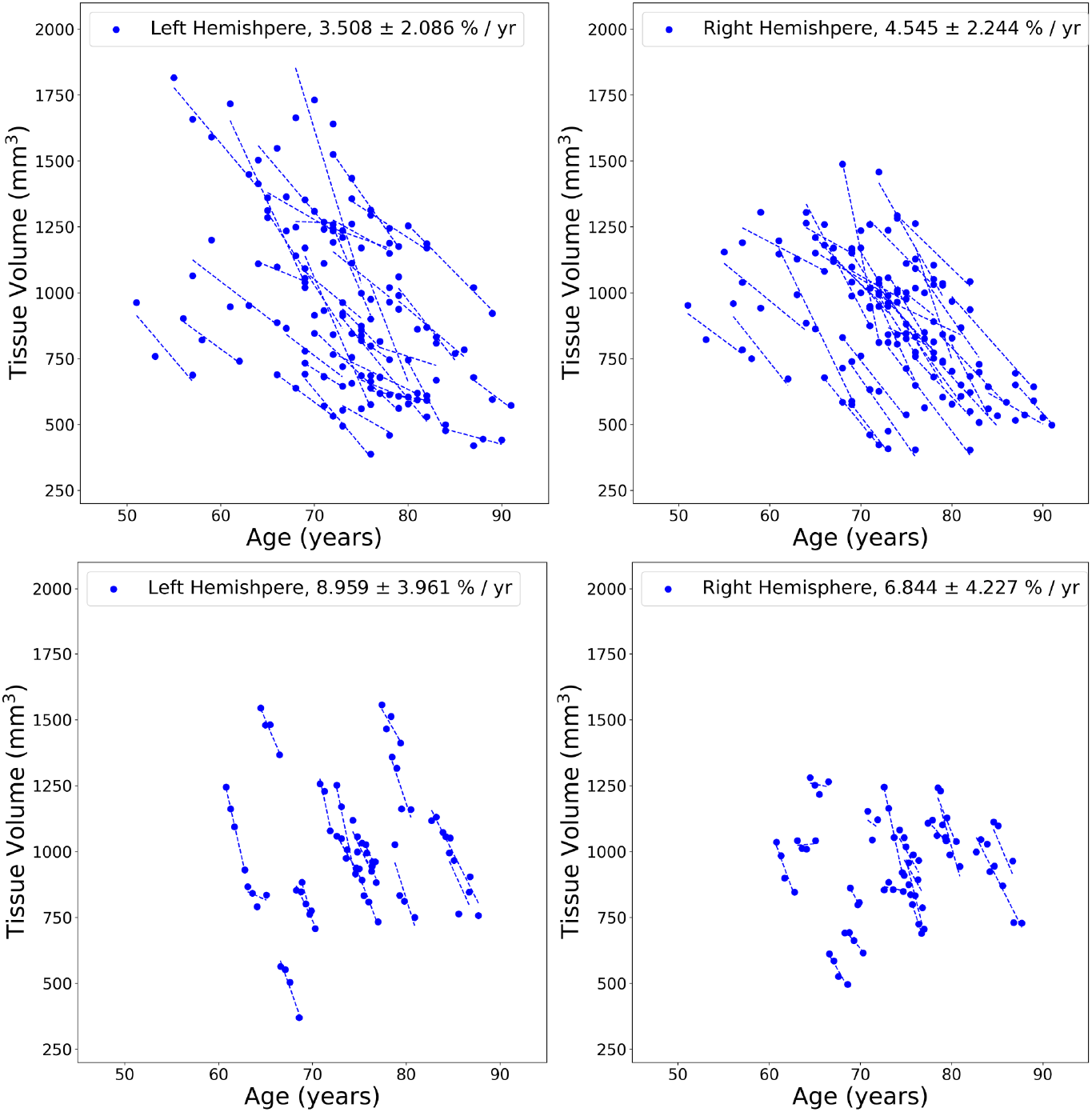
Volume atrophy rates in ERC/TEC (left and right sides represented by left and right columns respectively) for BIOCARD (top) and ADNI (bottom), MCI/AD group. Dots in each panel represent volume at certain age. A line is fitted through all timepoints for each subject.

Figures 7 and 8 show the summaries of the BIOCARD and ADNI cohorts in the MTL structures exhibiting the stereotyped monotonic behavior of ERC ∼ Amygdala *>* Hippocampus in controls, MCI and AD groups. This ordering is consistent between both left and right sides.

**Figure 7.**
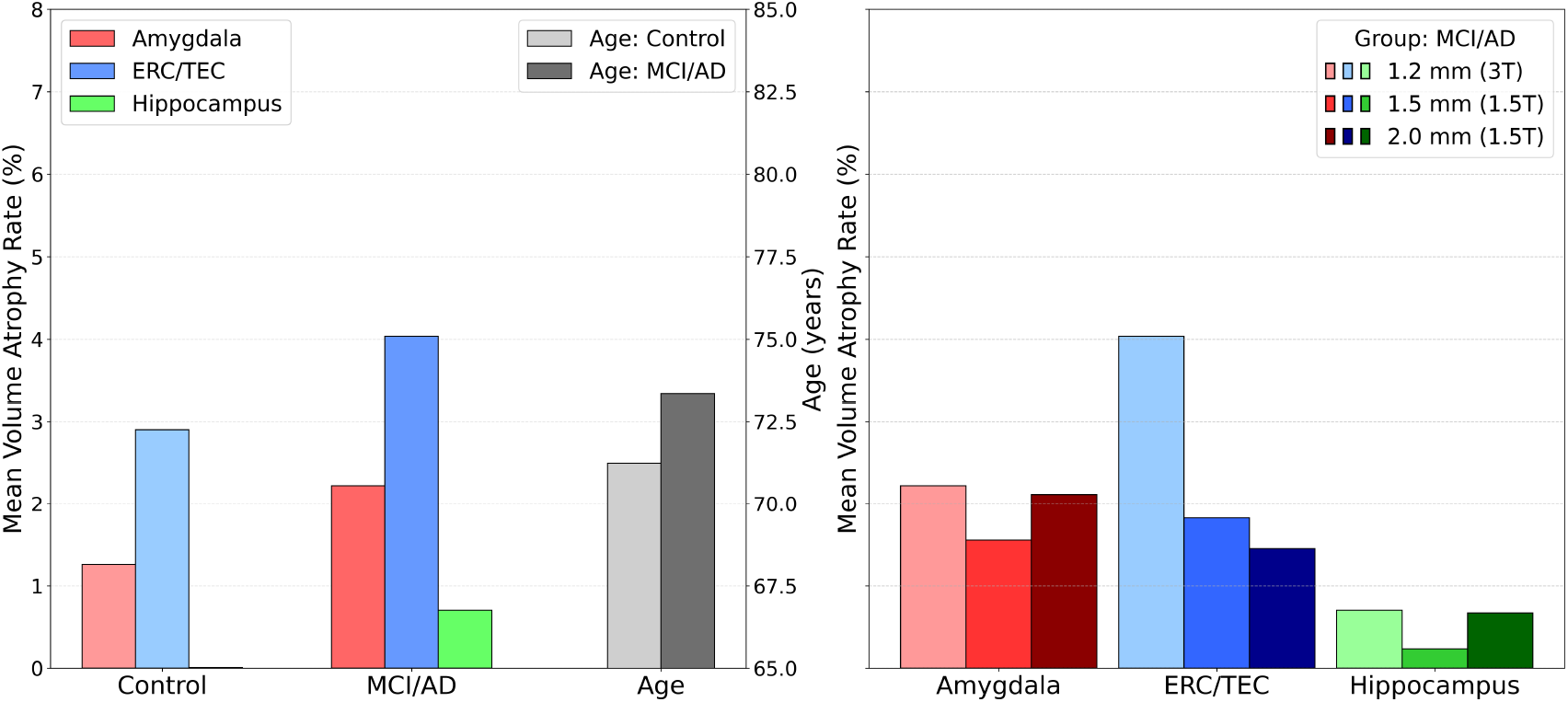
BIOCARD volume atrophy rates (left panel) of amygdala (red), ERC/TEC (blue), and hippocampus (green) with average ages for each disease group; volume atrophy rates for amygdala (red), ERC/TEC (blue), and hippocampus (green) for 1.5T and 3T scans.

**Figure 8.**
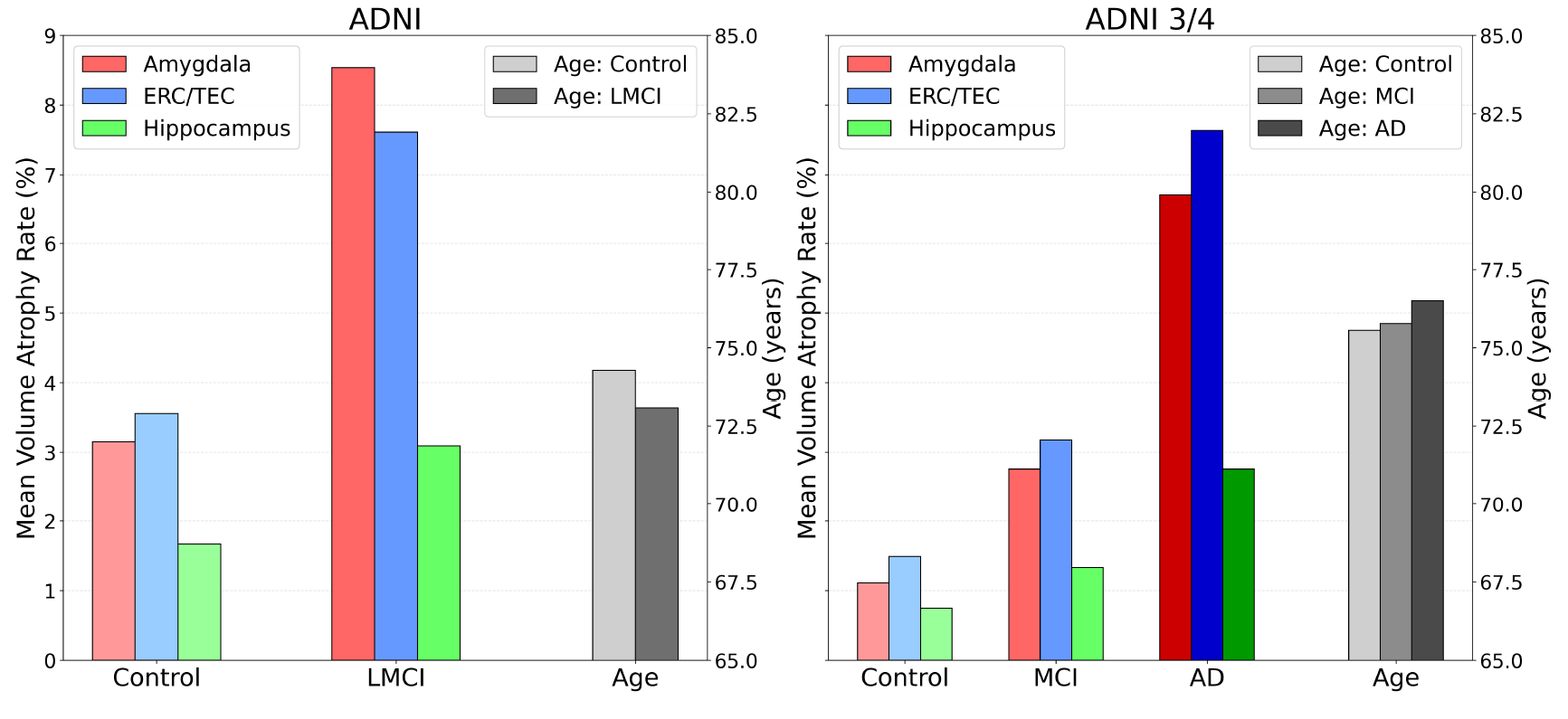
Volume atrophy rates ADNI 1, 2 & GO (left) and ADNI 3 & 4 (right) showing consistent ordering of atrophy: ERC ∼ Amygdala *>* Hippocampus; atrophy rates of amygdala (red), ERC/TEC (blue), and hippocampus (green). Average age of each disease group right in grey.

For BIOCARD and ADNI, the hippocampus atrophy is markedly lower. In BIOCARD (3T scans), interestingly the hippocampus atrophy rates are near zero at 0.01% and 0.70% in control and MCI groups, respectively. The amygdala atrophy increases dramatically in both groups to 1.26% and 2.22% in control and MCI, respectively. The ERC is increased as well to 2.90% and 4.03%. For ADNI (stage 1, 2 & GO) the hippocampus atrophy rates are 1.67% and 3.09% in control and MCI, respectively, with the amygdala 3.15% and 8.53% and the ERC 3.56% and 7.61%.

Subjects diagnosed with MCI or AD exhibit greatest atrophy in the entorhinal ERC/TEC with the highest rate of annualized volume loss, highlighting its role as an early and sensitive marker of AD-related neurodegeneration. Strikingly, the amygdala also demonstrated a remarkable early and consistent atrophy rate that is more consistent with the ERC far surpassing that of the hippocampus in early disease, reinforcing its emerging significance in the pathology of early-stage AD. This ordering is preserved across left and right hemispheres, and across cohort studies, suggesting a stereotyped progression of degeneration.

### 3.4. Volume atrophy in Entorhinal Cortex is linked to its laminar thickness atrophy rate

Figures 9 and 10 show the cortical surface reconstruction components. Top row of Figure 9 depicts the distribution of cortical thickness across a surface of a single individual. Middle row shows the density function of generated thickness and bottom row shows the cumulative distribution function for the time series of one individual in control and MCI groups, respectively. Atrophy rates of cortical thickness is obtained as the 50% points of the CDFs. Figure 10 depicts the collages of surfaces from subsets of ADNI (left) and BIOCARD (right).

**Figure 9.**
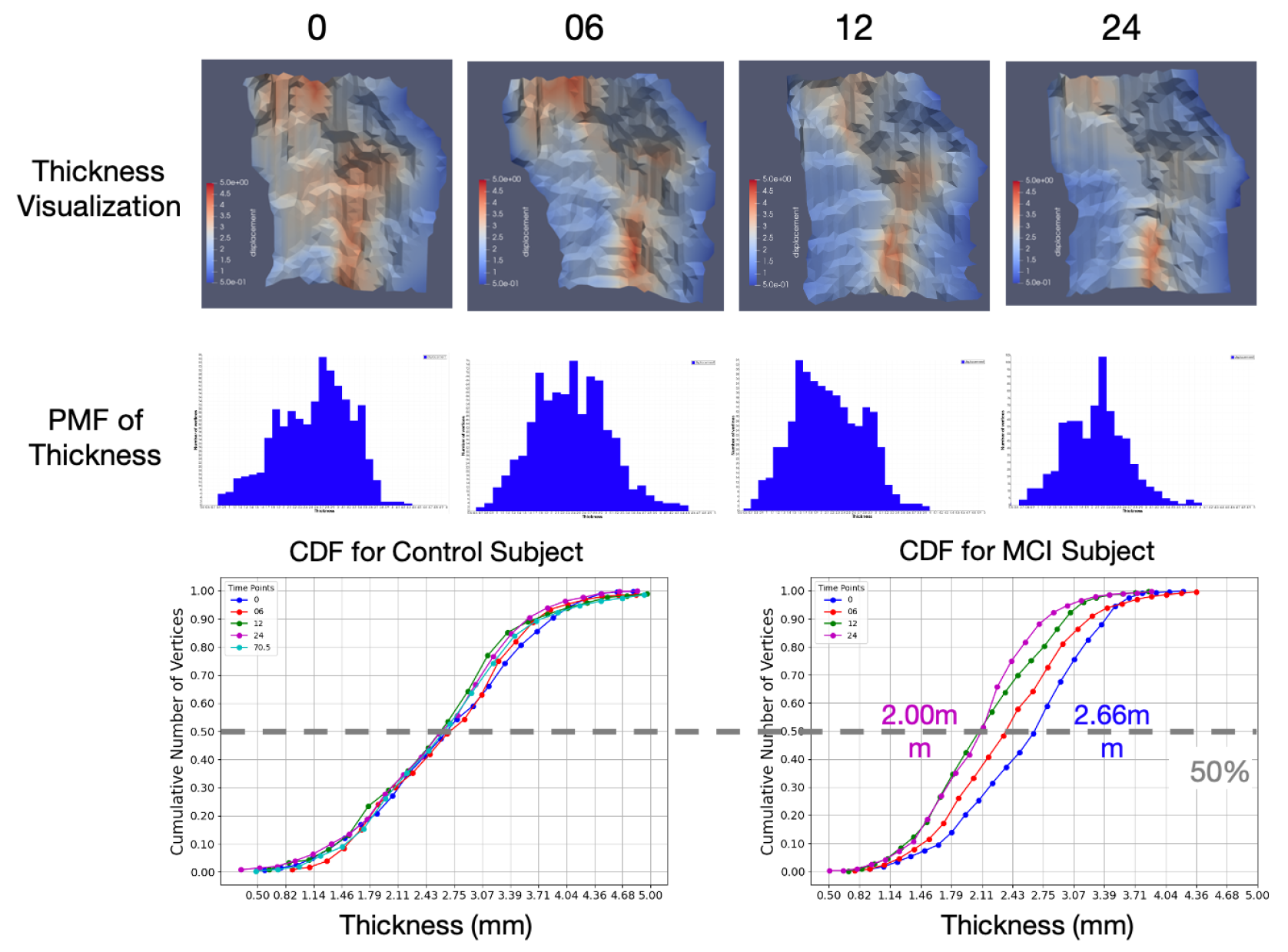
Top: cortical thickness across ERC/TEC surfaces at four time points for a single individual. Middle: corresponding probability mass (PMF) of thickness. Bottom: typical CDFs of subjects in control and MCI groups.

**Figure 10.**
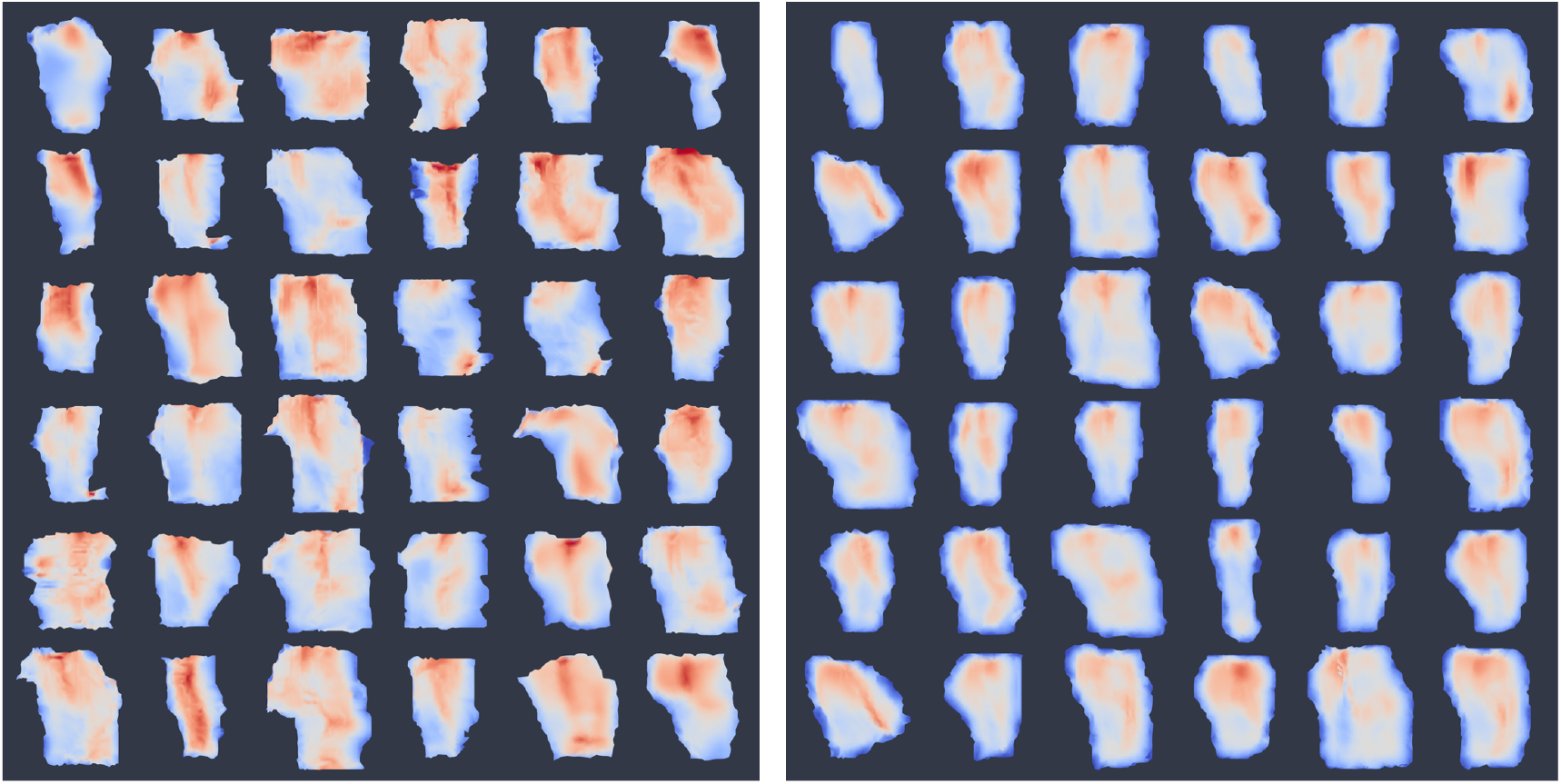
Collage of thickness (on a scale of 1–5 mm) on inner surface of ERC/TEC from subsets of ADNI (left) and BIOCARD (right) subjects.

Consistent with the volumetric trends, vertex-wise cortical thickness mapping of the ERC/TEC surface further revealed a graded spatial pattern of atrophy across clinical stages. Thickness is quantified by histogram analysis of vertex-wise annualized atrophy rates. Control subjects in Figure 11 shows lightly clustered, near-zero atrophy rate, whereas MCI subjects exhibited a heavy-tailed distribution with elevated atrophy, indicating more extensive and accelerated degeneration. This progression in both spatial extent and statistical distribution of atrophy further underscores the ERC/TEC region’s sensitivity to early AD-related neurodegeneration.

**Figure 11.**
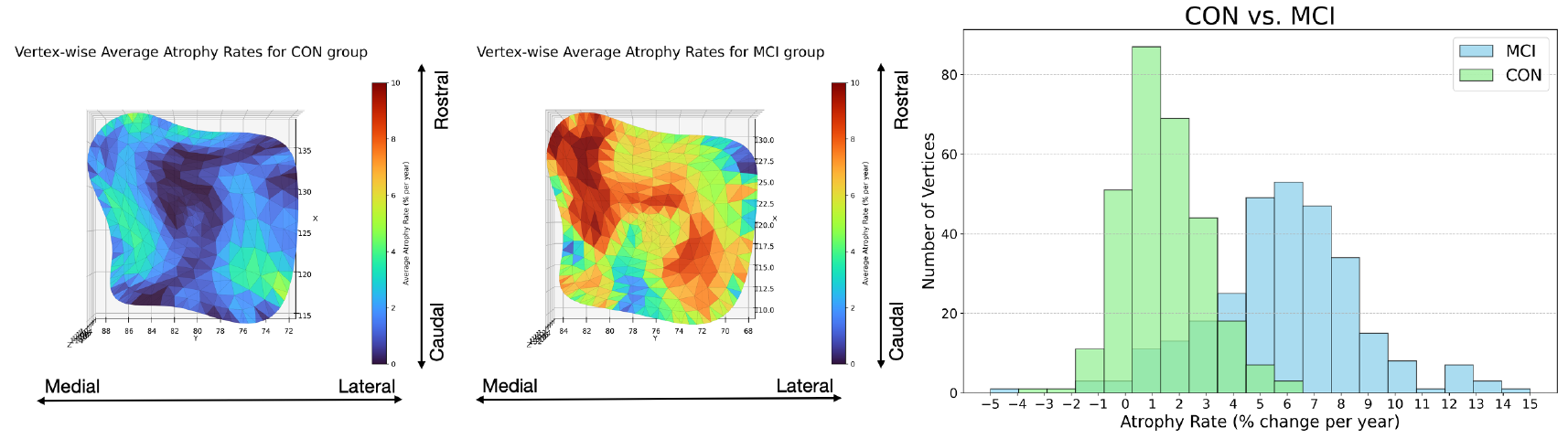
Averaged vertex-wise thickness atrophy rates: superimposed on baseline population outer surface (left and middle respectively for CON and MCI groups) and histogram (right) for both groups.

To explore the anatomical underpinning of volume atrophy in ERC/TEC, we examined the relationship between volume loss and cortical thinning. The top row of Figure 12 shows cortical thickness reconstruction from a subject at four time points. From these reconstructions, atrophy rates were calculated across the populations. The bottom row of Figure 12 depicts volume atrophy rates (y-axis) versus cortical thickness atrophy rates (x-axis) for the cohort of subjects in BIOCARD (left panel) and ADNI (right panel). Regression analysis indicates a linear correlation between ERC/TEC volume atrophy rate and cortical thickness loss, as derived from surface displacement measurements. In BIOCARD and ADNI MCI/AD subjects, the volumetric reductions are closely tied to laminar cortical surface thickness as manifest across the geometry of the surface. These results imply that ERC/TEC atrophy measured through volumetrics mirrors underlying laminar deterioration, affirming the biological validity of these imaging biomarkers. Such multi-dimensional confirmation strengthens the argument for ERC as a sentinel region in the early neurodegenerative cascade.

**Figure 12.**
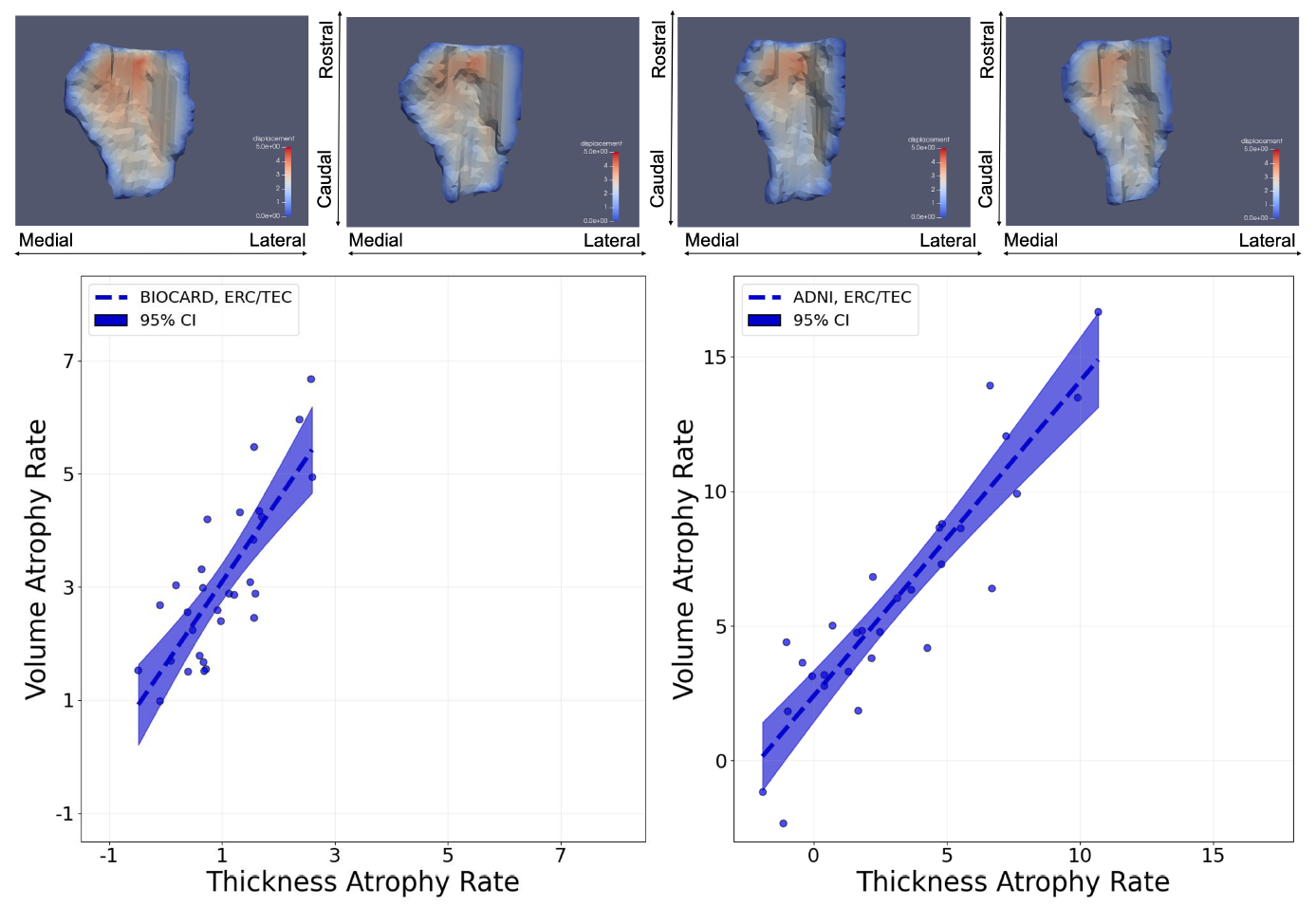
Top: thickness on inner surfaces of ERC/TEC rendered from segmentations from a BIOCARD at four time points. Bottom: volume atrophy rate versus thickness atrophy rate; BIOCARD (left) and ADNI (right).

### 3.5. Subregional Amygdalar Atrophy coupled to High Field Atlasing

To resolve the spatial specificity of atrophy within the amygdala, we conducted subregional analyses using atlas-based deformations involving high field amygdala reconstructions into the four major regions: BMA, BLA, CMA, and LA. These are shown in Figure 13. The MRI volumes were then mapped onto the four volume regions with atrophy rates calculated for each subregion. Volume loss was found to be notably heterogeneous across amygdala subnuclei. The BLA, BMA and CMA regions exhibited the greatest atrophy rates in MCI subjects, while lateral amygdala was comparatively spared. Interestingly, these findings mirror known patterns of tau pathology observed in histological studies and align with the amygdala’s structural and functional connectivity to other MTL regions such as the ERC and hippocampus.

**Figure 13.**
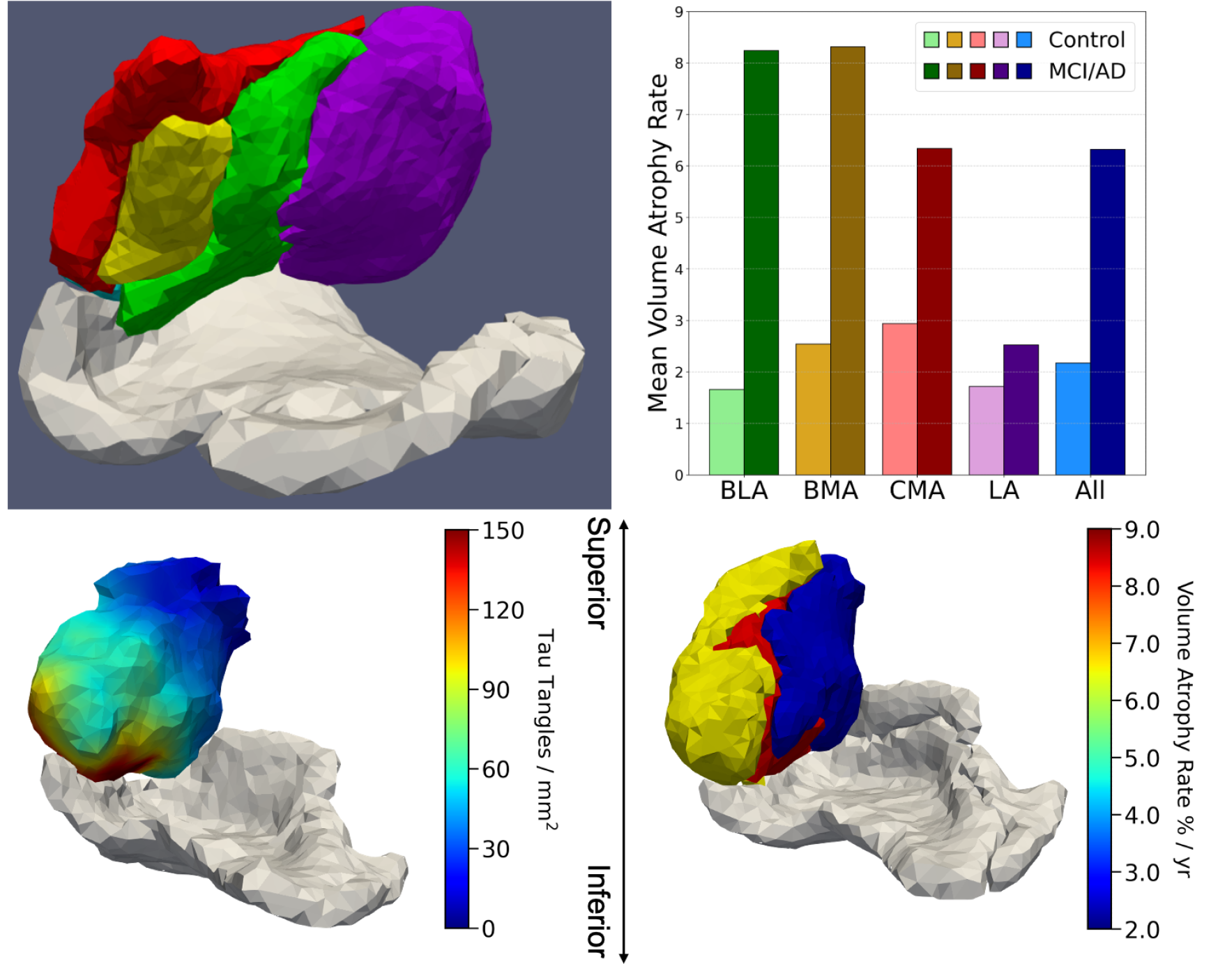
Top: Left shows subregions of amygdala including basomedial amygdala (BMA), basolateral amygdala (BLA), corticocentromedial amygdala (CMA), lateral amygdala (LA), and periamygdaloid area (PA); green BLA, yellow BMA, red CMA, purple LA. Right shows volume atrophy rates of subregions in deformed high-field atlas in Control (light) and MCI (dark) for the ADNI cohort. Bottom: Left shows NFT densities of amygdala in an AD postmortem sample [12] including BMA, BLA, CMA, LA; outlines of ERC and TEC surfaces shown in white mesh. Right shows corresponding average volume atrophy rates of subregions in deformed high-field atlas in the MCI/AD group from ADNI

The bottom row of Figure 13 demonstrates that atrophy in the amygdala based on the high-field atlases is consistent with the actual tau profiles as reconstructed by Stouffer et al. [12] in the high field digital pathology integration of the tau misfolded protein. Also shown are neurofibrillary tangle (NFT) densities of amygdala in the AD postmortem sample including BMA, BLA, CMA, and LA. The white mesh depicts the outlines of ERC and TEC surfaces shown. The right panels shows corresponding average volume atrophy rates of subregions in deformed high-field atlas in the MCI/AD group from ADNI cohort. Notice that the dark blue of the atrophy marker is at the identical LA lateral side of the amygdala as the tau density profile.

### 3.6. The MRI atrophy measures are correlated to rostral disease and are associated with tau burden as demonstrated by the high-field atlasing and tau PET imaging

Figure 14 illustrates a section of the rostral extent of the tauopathy and MRI atrophy within the epicenter of the high field digital pathology NFT tangles as well as the localized atrophy as measured via the MRI high field atlas mapping. Also shown are the volume atrophy rates for the BMA (8.24%), BLA (8.31%), CMA (6.34%), and LA (2.52%) subregions in deformed high-field atlas in the MCI/AD group from ADNI cohort. Notice that the dark blue decrease in the NFT density profile (left panel) is at the identical LA lateral side as the decreased atrophy marker in the LA amygdala (right panel).

**Figure 14.**
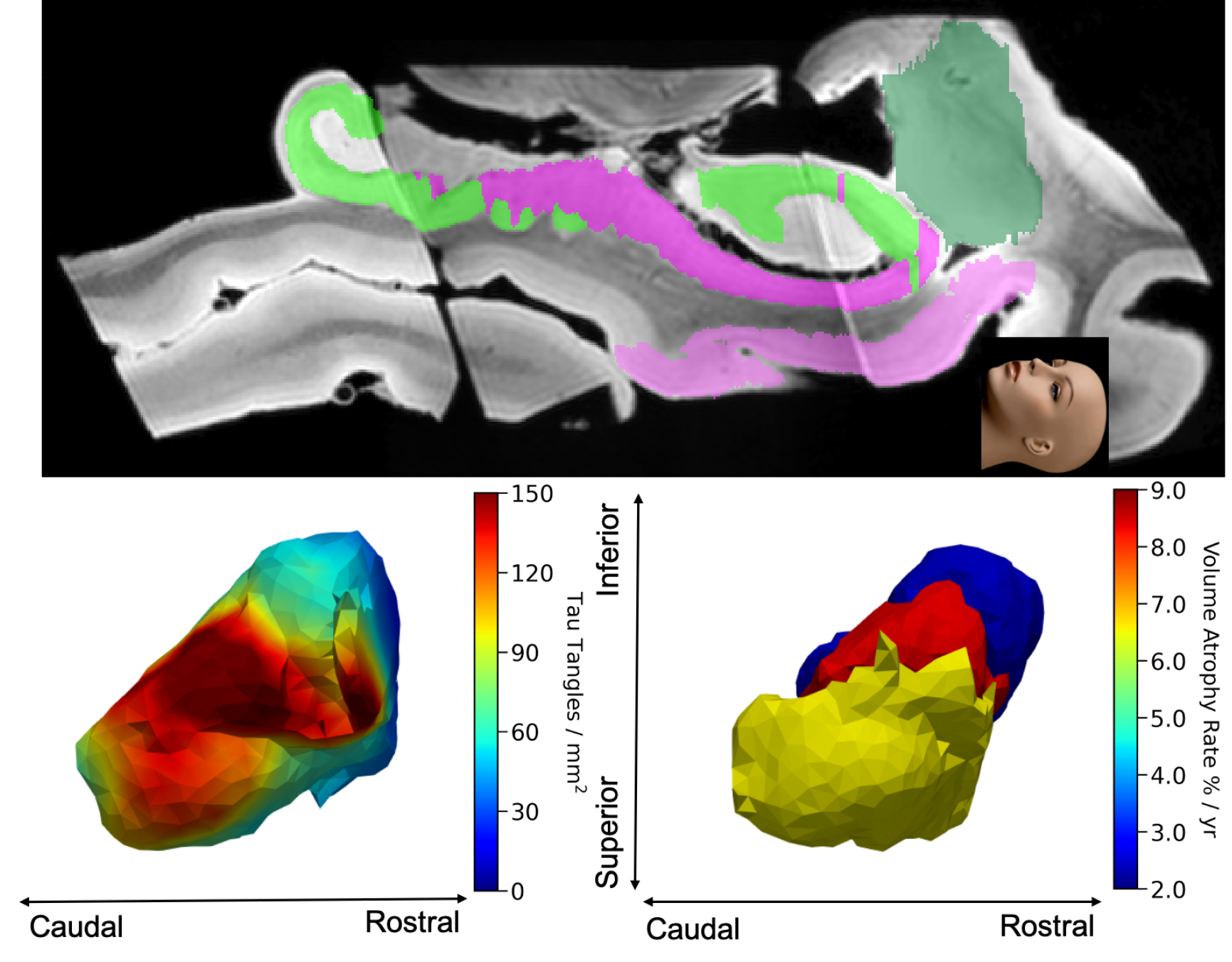
Top: a section through the high field MRI depicting the caudal (left) to rostral extent (right) of the amygdala (dark green) and its position relative to the rostral extent of the entorhinal cortex (light pink). Bottom: the tau NFTs (left) of the amygdala looking superiorly from the entorhinal cortex (avatar face looking up). The right panel shows volume atrophy rates of the MRI atrophy marker (right) BMA, BLA, CMA, and LA regions in the deformed high-field atlas in the MCI/AD group from ADNI cohort.

Figure 15 examines the relationship between the region-specific atrophy markers and their association to tau burden as measured on PET scans. The left column shows the tau markers measured in the BIOCARD (top row) and ADNI populations (bottom row). Tau PET accumulation (left column) is plotted versus the straight line *y* = *x* exhibiting strong inter-hemispheric symmetry. The fact that green points for the hippocampus are predominantly in the lower left part of the *y* = *x* with the ERC in the upper right quadrant demonstrates the delayed atrophy progression of the disease in the hippocampus relative to the ERC. This bilateral consistency also suggests that the PET-derived biomarkers are stable and robust across hemispheres, enhancing confidence in their use for individual subject-level tracking. The consistency served as a further validation of our segmentation, volume quantification, and PET normalization procedures resulting from the alignment pipeline of tau images to the segmented MRI volume. The right two columns demonstrate the relationship between tau PET accumulation and volume atrophy within the ERC/TEC (middle) and hippocampus (right) in the MCI/AD subjects. Regression lines fitting through individual data points reveal consistent trends across regions, underscoring the fact that the atrophy marker is highly correlated with tau PET accumulation and is region specific. Elevated tau accumulation, as measured via PET imaging, strongly predict accelerated volume loss in corresponding anatomical regions.

**Figure 15.**
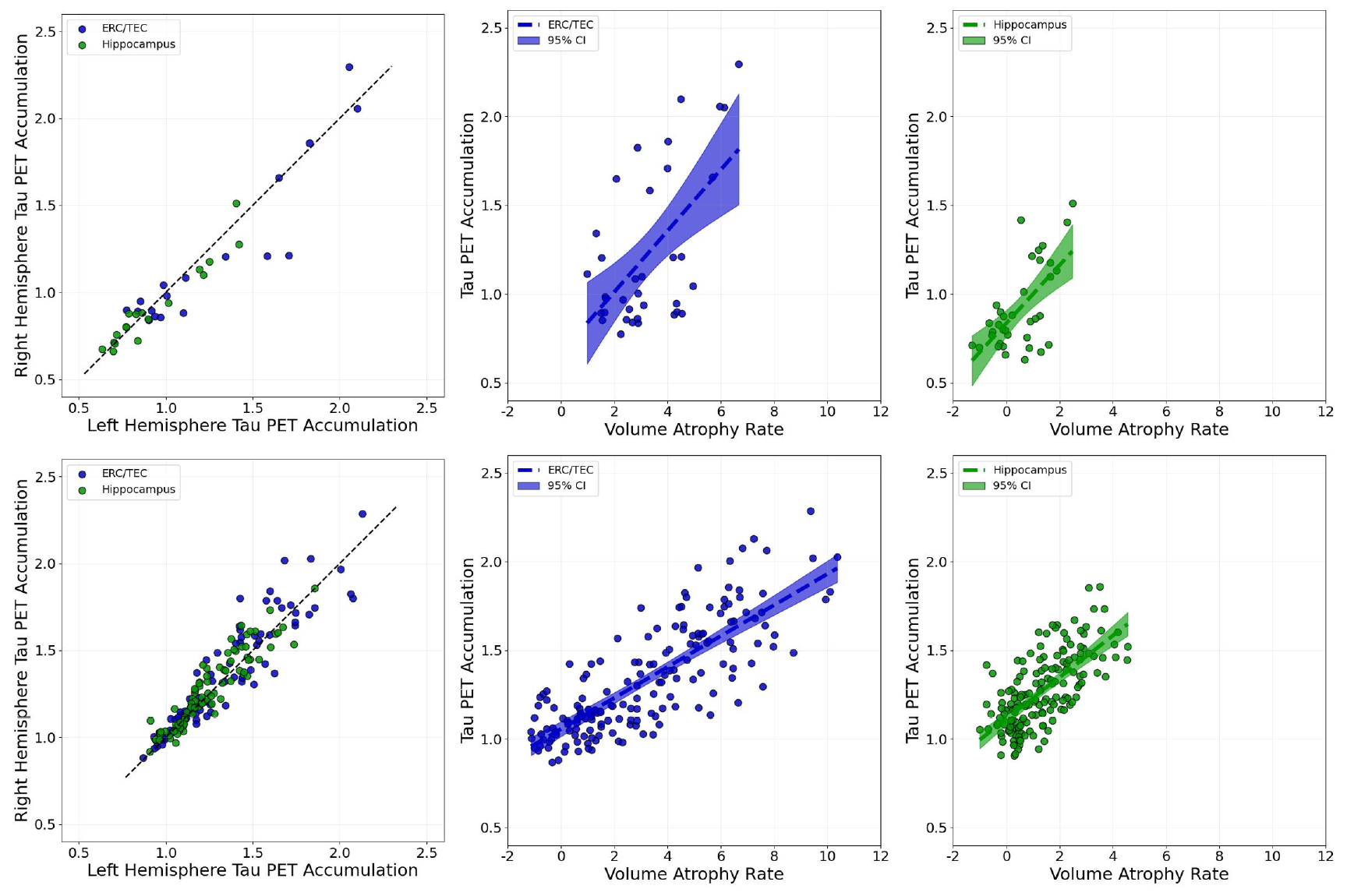
Left column shows tau PET consistency of the ERC (blue) and hippocampus (green); left structures plotted on *x*-axis and right structures on *y*-axis; straight line is the *y* = *x* linear trend. Middle and right columns show the linear regression of tau PET accumulation (*y*-axis) versus volume atrophy (*x*-axis) in the ERC (middle column, blue) and hippocampus (right column, green), with each group shown with shaded area denoting the 95% Confidence Interval (CI). Top and bottom rows respectively show the BIOCARD and ADNI 3 & 4 cohorts for MCI/AD subjects.

## 4. Discussion

This paper explores quantifiable medial temporal lobe atrophy associated to specific regions implicated in Alzheimer’s disease. While the hippocampus has historically been emphasized in the study of Alzheimer’s disease-related neurodegeneration, the amygdala and entorhinal cortex demonstrate greater atrophy of laminar thickness (ERC) and volume (amygdala) than the hippocampus.

Additionally, we quantified local cortical thinning in the ERC in the context of earlier work linking MRI-based atrophy with hyperphosphorylated tau deposition, deposition, showing our cortical thinning metrics to correspond to the layer specific deposition of tau. These findings align with neuropathological studies identifying tau burden as a primary driver of neuronal loss and regional atrophy in AD. The ability to detect subfield-specific atrophy in vivo holds promise for improving sensitivity and specificity in early AD diagnosis and monitoring.

Interestingly, while the amygdala plays a central role as an interconnected hub to critical brain regions in the memory circuit and is clearly an early actor exhibiting pathologic changes during neurodegeneration, relatively less is known about the molecular and cellular level changes in the amygdala in AD. The amygdala innervates many of the brain structures crucially affected by early tau pathology in AD, including the ERC, hippocampus, basal forebrain nuclei, and olfactory bulb [4, 55]. Experimental evidence implicates the amygdala in fear processing, anxiety, aggression, stress, motivation, feeding behaviors, and depression [56, 57, 58]. While not central to the classic symptomatology of AD, these behaviors constitute many of the diagnostic criteria for mild behavioral impairment (MBI), which is emerging as an early clinical marker of AD [59] and consequently a support for a role of the amygdala in early AD. Additionally, there is evidence that the amygdala plays a role in long term memory modulation, particularly memories associated with strong emotion, and in working memory and attention [57], as traditionally implicated in the AD spectrum.

The functional and connective differences in subregions of the amygdala provide an interesting window into the possible circuit-level changes affected by the spatially-distributed pathology in the amygdala in AD and related dementia. Our studies here and others [8, 9, 12] have demonstrate that tau pathology in the amygdala in AD is distributed in specific spatial patterns. Generally, studies of postmortem human tissue in AD agree that the more lateral regions in the amygdala have less tau pathology compared to the more medial regions [8, 9, 12, 60], though the anatomic definitions of regions vary. At a circuit-level, these medial subregions are shown to be linked to the anterior hippocampus, ERC, and locus ceruleus [58], suggesting possible neuronal links for spread of pathologic tau in AD [61] between each of these historically implicated areas. In future, improved understanding of the sub-regional and cell-type specific changes in the amygdala could provide insight into the potential presence of multiple circuits of tau spread through the medial temporal lobe and the circuit-level dysfunction associated with cognitive and neuropsychiatric symptoms in AD.

## Data Availability

The data used in this study are available from the Alzheimer's Disease Neuroimaging Initiative (ADNI) database (https://adni.loni.usc.edu
) and the BIOCARD study at Johns Hopkins University, subject to data use agreements and application approval. Derived data supporting the findings of this study are available from the corresponding author upon reasonable request.

## Credit Author Statement

**Michael I. Miller**: Conceptualization, Methodology, Formal analysis, Writing - original draft, Writing - review & editing, Supervision, Project administration, Funding acquisition. **Yi Xie**: Conceptualization, Methodology, Formal analysis, Investigation, Data curation, Software, Validation, Visualization, Writing - original draft, Writing - review & editing. **Kaitlin M. Stouffer**: Conceptualization, Methodology, Data curation, Formal analysis, Writing - review & editing. **Can Ceritoglu**: Data curation, Software, Writing - review & editing. **Jiabei Li**: Data curation. **J. Tilak Ratnanather**: Writing - review & editing. **Laurent Younes**: Conceptualization, Methodology, Formal analysis, Writing - review & editing. **Arnold Bakker**: Data curation, Writing - review & editing. **Nisha Rani**: Data curation. **Marilyn Albert**: Writing - review & editing, Funding acquisition. **Juan Troncoso**: Conceptualization, Formal analysis, Resources, Data curation, Writing - review & editing. **Meaghan Morris**: Conceptualization, Formal analysis, Resources, Data curation, Writing - review & editing.

## Acknowledgements

Data from the ADNI study was funded by National Institutes of Health Grant U01 AG024904 and DOD ADNI (Department of Defense award number W81XWH-12-2-0012). ADNI is funded by the National Institute on Aging, the National Institute of Biomedical Imaging and Bioengineering, and through generous contributions from the following: AbbVie, Alzheimer’s Association; Alzheimer’s Drug Discovery Foundation; Araclon Biotech; BioClinica, Inc.; Biogen; Bristol-Myers Squibb Company; CereSpir, Inc.; Eisai Inc.; Cogstate; Elan Pharmaceuticals, Inc.; Eli Lilly and Company; EuroImmun; F. Hoffmann-La Roche Ltd and its affiliated company Genentech, Inc.; Fujirebio; GE Healthcare; IXICO Ltd.; Janssen Alzheimer Immunotherapy Research & Development, LLC.; Johnson & Johnson Pharmaceutical Research & Development LLC.; Lumosity; Lundbeck; Merck & Co., Inc.; Meso Scale Diagnostics, LLC.; NeuroRx Research; Neurotrack Technologies; Novartis Pharmaceuticals Corporation; Pfizer Inc.; Piramal Imaging; Servier; Takeda Pharmaceutical Company; and Transition Therapeutics. The Canadian Institutes of Health Research is providing funds to support ADNI clinical sites in Canada. Private sector contributions are facilitated by the Foundation for the National Institutes of Health (www.fnih.org). The grantee organization is the Northern California Institute for Research and Education, and the study is coordinated by the Alzheimer’s Therapeutic Research Institute at the University of Southern California. ADNI data are disseminated by the Laboratory for Neuro Imaging at the University of Southern California.

Data from the BIOCARD study are supported by grant U19 AG033655 from the National Institute on Aging. The BIOCARD Study consists of 7 Cores with the following members: (1) The Administrative Core (MA, Barbara Rodzon), (2) the Clinical Core (MA, Anja Soldan, Rebecca Gottesman, Ned Sacktor, Corinne Pettigrew, Scott Turner, Leonie Farrington, Maura Grega, Jules, Gilles, Gay Rudow, Rostislav Brichko, Scott Rudow), (3) the Imaging Core (MM, Susumu Mori, Tilak Ratnanather, Anthony Kolasny, Hanzhang Lu, Kenichi Oishi, LY), (4) the Biospecimen Core (Abhay Moghekar, Jacqueline Darrow, Richard O’Brien), (5) the Informatics Core (Roberta Scherer, David Shade, Ann Ervin, Jennifer Jones, Hamadou Coulibaly, Kathy Moser), the (6) Biostatistics Core (Mei-Cheng Wang, Yuxin Zhu, Jiangxia Wang), and (7) the Neuropathology Core (Juan Troncoso, Olga Pletnikova, Karen Fisher). We would also like to acknowledge the members of the BIOCARD Scientific Advisory Board who provide oversight an guidance regarding the conduct of the study including: Drs. John Csernansky, David Holtzman, David Knopman, Walter Kukull and Kevin Duff, as well as Drs. Laurie Ryan and John Hsiao, who provide oversight on behalf of the NIA. Additionally, we recognize the members of the BIOCARD Resource Allocation Committee who provide ongoing guidance regarding the use of the biospecimens collected as part of the study, including: Drs. Constantine Lyketsos, Carlos Pardo, Gerard Schellenberg, Leslie Shaw, Madhav Thambisetty, and John Trojanowski. We would like to acknowledge the contributions of the Geriatric Psychiatry Branch (GPB) of the intramural program of the NIMH who initiated this study (PI: Dr. Trey Sunderland). Dr. Karen Putnam provided documentation of the GPB study procedures and the datafiles received from NIMH. We acknowledge the altruism of the participants and their families and contributions of the BIOCARD research and support staff for their contributions to this study. We thank Henry Noren for his work organizing data and running software. We thank Ophelia Yang for her work on augmenting training data for machine learning. We thank Fabiana Amato for her work analyzing manual segmentations.

